# The RBD Of The Spike Protein Of SARS-Group Coronaviruses Is A Highly Specific Target Of SARS-CoV-2 Antibodies But Not Other Pathogenic Human and Animal Coronavirus Antibodies

**DOI:** 10.1101/2020.05.06.20093377

**Authors:** Lakshmanane Premkumar, Bruno Segovia-Chumbez, Ramesh Jadi, David R. Martinez, Rajendra Raut, Alena Markmann, Caleb Cornaby, Luther Bartelt, Susan Weiss, Yara Park, Caitlin E. Edwards, Eric Weimer, Erin M. Scherer, Nadine Roupael, Sri Edupuganti, Daniela Weiskopf, Longping V. Tse, Yixuan J. Hou, David Margolis, Alessandro Sette, Matthew H. Collins, John Schmitz, Ralph S. Baric, Aravinda M. de Silva

**Affiliations:** Department of Microbiology and Immunology, University of North Carolina School of Medicine, Chapel Hill NC 27599, USA; Department of Epidemiology, UNC Chapel Hill School of Public Health, University of North Carolina at Chapel Hill, Chapel Hill, North Carolina, USA; Departments of Medicine, University of North Carolina School of Medicine, Chapel Hill NC 27599, USA; Department of Pathology & Laboratory Medicine, University of North Carolina School of Medicine, Chapel Hill NC 27599, USA; Immunology/Histocompatibility and Immunogenetics Laboratories, University of North Carolina School of Medicine, Chapel Hill NC 27599, USA; Center for Infectious Disease and Vaccine Research, La Jolla Institute for Immunology (LJI), La Jolla, CA 92037, USA; Department of Medicine, Division of Infectious Diseases and Global Public Health, University of California, San Diego (UCSD), La Jolla, CA 92037, USA; Hope Clinic of the Emory Vaccine Center, Division of Infectious Diseases, Department of Medicine, School of Medicine, Emory University, Decatur, Georgia, USA

**Author notes:** Correspondence should be addressed to Lakshmanane Premkumar or Aravinda M. de Silva.

## Abstract

A new Severe Acute Respiratory Syndrome Coronavirus variant (SARS-CoV-2) that first emerged in late 2019 is responsible for a pandemic of severe respiratory illness. People infected with this highly contagious virus present with clinically inapparent, mild or severe disease. Currently, the presence of the virus in individual patients and at the population level is being monitored by testing symptomatic cases by PCR for the presence of viral RNA. There is an urgent need for SARS-CoV-2 serologic tests to identify all infected individuals, irrespective of clinical symptoms, to conduct surveillance and implement strategies to contain spread. As the receptor binding domain (RBD) of the viral spike (S) protein is poorly conserved between SARS-CoVs and other pathogenic human coronaviruses, the RBD represents a promising antigen for detecting CoV specific antibodies in people. Here we use a large panel of human sera (70 SARS-CoV-2 patients and 71 control subjects) and hyperimmune sera from animals exposed to zoonotic CoVs to evaluate the performance of the RBD as an antigen for accurate detection of SARS-CoV-2-specific antibodies. By day 9 after the onset of symptoms, the recombinant SARS-CoV-2 RBD antigen was highly sensitive (98%) and specific (100%) to antibodies induced by SARS-CoVs. We observed a robust correlation between levels of RBD binding antibodies and SARS-CoV-2 neutralizing antibodies in patients. Our results, which reveal the early kinetics of SARS-CoV-2 antibody responses, strongly support the use of RBD-based antibody assays for population-level surveillance and as a correlate of neutralizing antibody levels in people who have recovered from SARS-CoV-2 infections.

## Introduction

A novel severe acute respiratory syndrome coronavirus (SARS-CoV-2) is responsible for an ongoing pandemic that has already killed over 250,000 people and paralyzed the global economy (1). Currently, the main method for laboratory diagnosis of SARS-CoV-2 is PCR testing of nasopharyngeal swabs. There is an urgent need for highly specific and sensitive antibody detection assays to answer fundamental questions about the epidemiology and pathogenesis of SARS-CoV-2 and to implement and evaluate population-level control programs (2). Efforts to understand the pathogenesis and define risk factors for severe SARS-CoV-2 disease have been hampered by our inability to identify all infected individuals, irrespective of clinical symptoms. To contain the pandemic, countries have resorted to the widespread quarantine of cities and regions, which has led to devastating social and economic crisis. By deploying reliable antibody assays for population-level testing, it will be possible to obtain the high-resolution spatial data needed to implement policies for containing the epidemic and informing strategies for re-opening communities and cities.

Studies with SARS-CoV-2 and other human CoVs demonstrate that people rarely develop specific antibodies within the first 7 days after onset of symptoms (3-7). By 10-11 days after onset of symptoms, greater than 90% of SARS-CoV-2 patients develop specific IgG and IgM (3-6). For SARS-CoV-1 and the more distantly related MERS-CoV, IgG antibodies have been observed to persist for at least one year after infection (8, 9). These observations strongly support the feasibility of using antibody assays for identifying recent and remote SARS-CoV-2 infections and for conducting population-level surveillance.

SARS-CoV-2 is a β-coronavirus that includes the closely related SARS-CoV-1 and the more distantly related MERS CoV and the common-cold human CoVs (HCoV-OC43 and HCoV-HKU1) (10). Many companies have quickly developed tests for SARS-CoV-2 antibody detection. These assays utilize the inactivated whole virion, viral nucleocapsid protein or viral spike protein as antigens in ELISA, lateral flow or other testing platforms. While the performance of these assays has not been fully evaluated, some assays appear quite sensitive when used 10 days or more after the onset of symptoms (6, 11). The specificity of SARS-CoV-2 antibody assays have not been adequately addressed. Humans are frequently infected with HCoV-OC43 and HCoV-HKU1 and most adults have antibodies to these viruses (10). Any antibody cross-reactivity between common HCoVs and SARS-CoV-2 would result in false-positives that will severely compromise antibody-based testing and surveillance for SARS-CoV-2.

SARS-CoV-1 and HCoV OC43 elicit antibodies that cross-react against related CoVs (12, 13). Following the SARS-CoV-1 outbreak in 2003, the overall specificity of serological assays utilizing the nucleocapsid protein of SARS-CoV-1 was poor, whereas assays based on the Spike protein were more specific (14-16). In recent studies, the receptor binding domain (RBD) of the spike protein of SARS-CoV-2 has shown promise as an antigen for specific antibody detection (4, 17, 18). Here we report the production of properly folded recombinant receptor binding domains (RBDs) from the spike proteins of SARS and common-cold HCoVs in mammalian cells. We use these recombinant antigens and a large diverse panel of human and animal sera to evaluate the RBD as an antigen for SARS-CoV-2 serology. We demonstrate that the recombinant SARS-CoV-2 RBD antigen is highly sensitive and specific to antibodies induced by SARS-CoVs. We also observed a strong correlation between the levels of RBD-binding antibodies and levels of SARS-CoV-2 neutralizing antibodies in patients. Our results provide strong support for the use of RBD-based antibody assays for population-level surveillance and as a correlate of neutralizing antibody levels in people who have recovered from SARS-CoV-2 infections.

## RESULTS

### Expression and Characterization of Recombinant RBD Antigens from Pathogenic Coronaviruses

The S1 and S2 subunits of the spike (S) protein of Coronaviruses is required for viral entry. The surface accessible receptor binding domain (RBD) on the S1 subunit binds to receptors on target cells, whereas the exposure of the fusion loop in the S2 subunit induces fusion of the viral envelope to the host cellular membranes (19). The RBD of SARS-CoVs, which binds to angiotensin-converting enzyme 2 (ACE2) receptor on the host cells, is also a major target of human antibodies (Figure 1A and B). As the RBD is a common target of human antibodies and poorly conserved between SARS-CoVs and other pathogenic human coronaviruses (Figure 1C), this domain is a promising candidate for use in antibody-based diagnostic assays. We expressed the RBD of 2003 and 2019 SARS-Co-Vs and four common human Coronaviruses (HCoV-HKU-1, -OC43, -NL63 and −229E) as fusion proteins that were secreted from human cells. The recombinant RBDs were purified from the cell culture medium by affinity chromatography and purity was confirmed by SDS-PAGE (Figure 1D). We used sera and monoclonal antibodies from animals immunized with SARS-CoV-1 or 2 spike proteins to assess the structural integrity of the purified recombinant RBD antigens. Pooled serum from mice immunized with SARS-CoV-2 spike protein had antibodies that bound well to the RBD of SARS-CoV-2 RBD and poorly to RBD of SARS-CoV-1 and other common HCoVs (Figure 1E). Sera from mice or rabbits immunized with SARS-CoV-1 or cross-reactive monoclonal antibody 240C reacted with the RBD of SARS CoV-1 and 2 but not common human CoVs (Figure 1E). Human serum collected before SARS-CoV-2 emerged contained antibodies to common α-and β-HCoVs but not to SARS CoV RBD antigens (Figure 1E). These results suggest that the purified recombinant RBD antigens retain native structures required for specific antibody binding.

**Figure 1.**
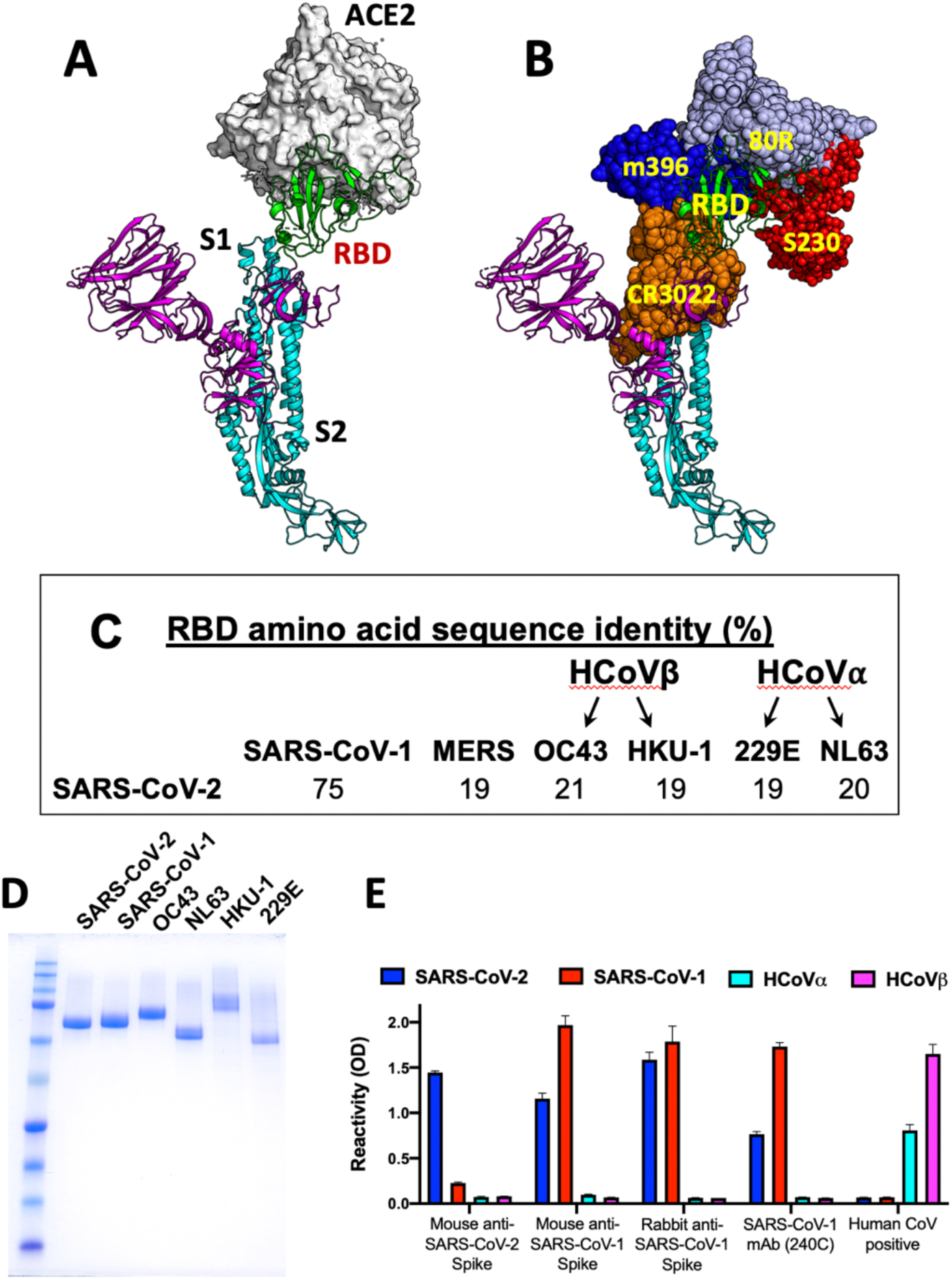
Production and characterization of the RBD of the Coronavirus spike antigens. **A**. The Spike protein on the virion surface engages its cognate receptor via the RBD. **B**. RBD of the spike protein is the main human Ab target in SARS-CoV-1. **C**. The amino acid sequence corresponding to RBD of the spike protein is poorly conserved between SARS-CoV-2 and common human coronaviruses. **D**. Coomassie-stained SDS-PAGE of purified spike RBD antigens from different CoVs. **E**. Binding characterization of the spike RBD antigens with immune sera and a monoclonal antibody. SARS-CoV-1 mAb (240C), serum from a mouse immunized with VRP expressing SARS-CoV-2 or SARS-CoV-1 spike protein, serum from a rabbit immunized with SARS-CoV-1 spike protein and an archived human sample collected before COVID-19 were tested for binding against RBD spike antigens from SARS-CoV-2, SARS-Co-V-1, HCoVα (NL63) and HCoVβ (HKU-1).

### Evaluating the specificity of SARS-CoV-2 RBD for serology

To evaluate the specificity of the recombinant SARS-CoV-2 RBD in serology, we used human sera collected from different populations before the current pandemic. The sera were tested at a high concentration (1:20 dilution) for binding to the recombinant RBDs from SARS-CoV-1, SARS-CoV-2 and common α- and β-HCoVs (Figure 2). Sera collected from healthy American adults (N = 20) before the SARS-CoV-2 pandemic frequently had high levels of antibodies to the recombinant RBDs of common α- and β-HCoVs but not to SARS-CoVs (Figure 2A). We also tested archived pre-SARS-CoV-2 pandemic sera collected from individuals in South Asia, the Caribbean and Central America who had recently recovered from arbovirus infections. As in the case of healthy adults from the USA, most of the subjects from different parts of the world had high levels of antibodies to the RBD of common HCoVs but no antibodies to the RBD of SARS-CoVs (Figure 2B). To assess if other human respiratory viruses stimulated antibodies that cross-reacted with the recombinant SARS-CoV RBD, we tested early convalescent sera from people with laboratory confirmed influenza A and respiratory syncytial virus infections and sera from guinea pigs immunized with a panel of different human respiratory viruses (Figure 2 C and D). Except guinea pigs immunized with SARS-CoV-1, none of the sera had detectable levels of antibodies to the recombinant RBD of SARS-CoVs.

**Figure 2.**
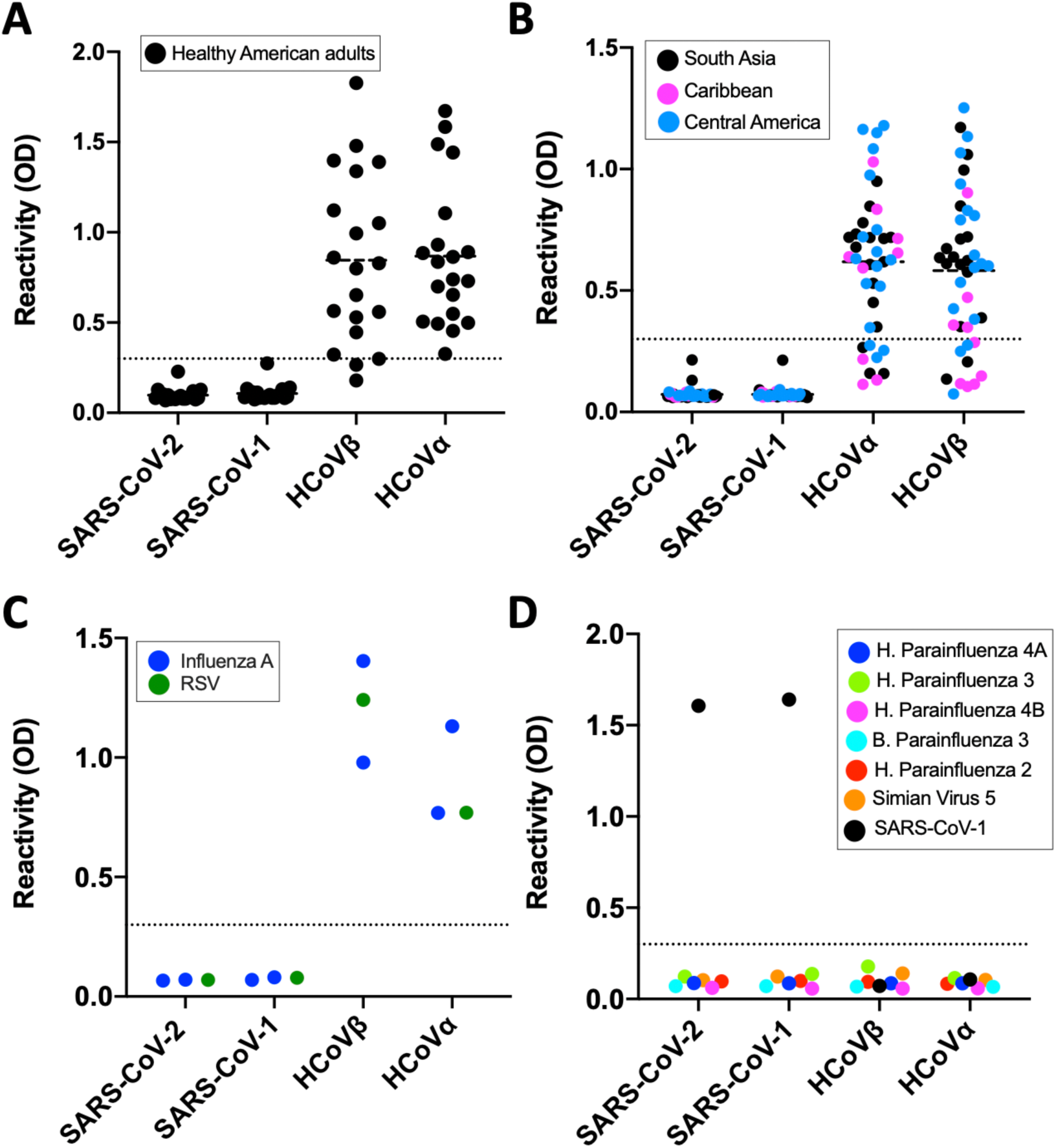
Evaluation of SARS-CoV-2 spike RBD antigen specificity using blood samples collected before the emergence of COVID-19. Spike RBD antigen binding was assessed by in-house ELISA assay against a panel of de-identified archived serum specimens obtained from **A**. American healthy adults, **B**. Convalescent sera from dengue/Zika patients in South Asia, Caribbean, and Central America, **C**. People who had recently recovered from viral respiratory illnesses, and **D**. Guinea pigs immunized with respiratory viruses or SARS-CoV-1 spike protein. The cutoff values for the ELISA assay are indicated by the broken line.

The known pathogenic human CoVs are members of the α-coronavirus and β-coronavirus genera (Figure 3A). HCoV-NL63 and 229E are two α-coronaviruses that frequently infect and cause a mild common-cold-like illness in most people. HCoV-OC43 and HKU-1 are two group 2A β-coronaviruses that also commonly infect people and cause mild disease. Most adults (>90%) have antibodies to these common-cold HCoVs. SARS-CoV-1 and 2 and MERS-CoV are group 2B and 2C zoonotic β-coronaviruses that have recently crossed into humans and caused severe illness. The α- and β-coronavirus genera also contain a large number of zoonotic viruses that infect different animal hosts, which have not been implicated in human disease to date. To further assess the specificity of SARS-CoV-2 RBD for serology, we obtained and tested sera from people who had recently recovered from a laboratory confirmed common-cold HCoVs infection and sera from guinea pigs immunized with different animal CoVs (Figure 3 B and C). None of the immune sera from people exposed to recent HCoV infections cross-reacted with the recombinant RBD of SARS-CoVs. None of the guinea pigs vaccinated with different zoonotic CoVs had antibodies that cross-reacted with the recombinant SARS-CoV RBDs (Figure 3B and C). These results establish that most individuals, including people who have been recently exposed to acute common HCoV infections, do not have detectable levels of cross-reactive antibodies to the recombinant RBD of SARS-CoVs.

**Figure 3.**
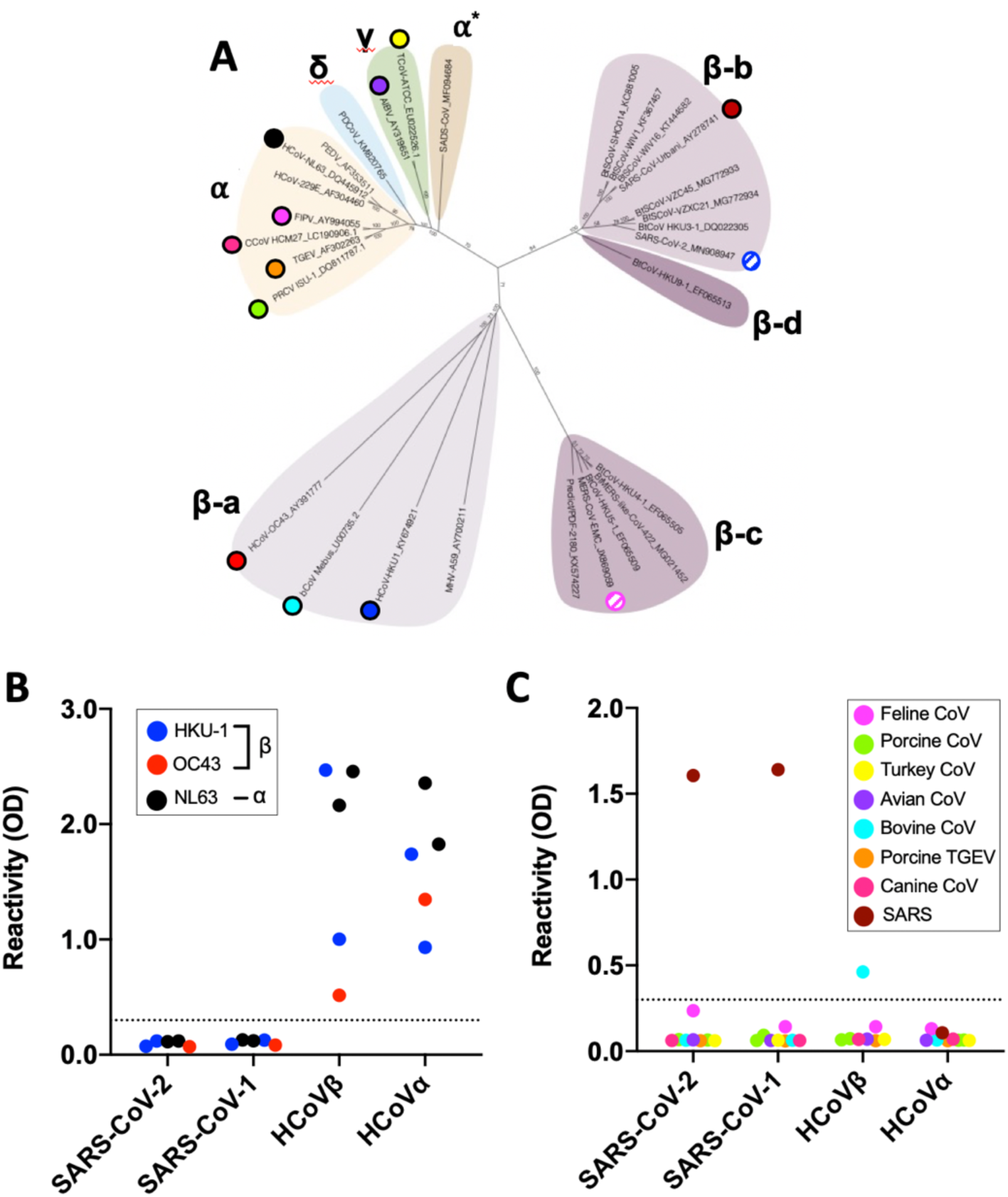
Evaluation of SARS-CoV-2 spike RBD antigen specificity against common human CoVs and animal CoVs sera. **A**. Phylogenic tree of the spike protein from representative coronaviruses. Coronavirus genera are grouped by classic subgroup designations (α, βa-d, γ, and δ). Numbers following the underscores in each sequence correspond to the GenBank Accession number. Spike RBD antigen binding was assessed by in-house ELISA assay using **B**. human convalescent samples obtained from PCR confirmed HCoVα (NL63, black) and HCoVβ (OC43 (red), HKU-1 (blue)) infections and **C**. sera from guinea pigs immunized with spike antigen from SARS-CoV-1 or indicated animal CoV. The cutoff values for the ELISA assay are indicated by the broken line. Feline Infectious Peritonitis Virus, 79-1146 (Feline CoV, Pink); respiratory coronavirus strain ISU-1(Porcine CoV, green); Porcine Transmissible Gastroenteritis Virus (TGEV, orange); Bovine Coronavirus strain mebus (Bovine CoV, cyan); Avian Infectious Bronchitis Virus, Massachusetts (Avian CoV, violet); Turkey Coronavirus, Indiana (Turkey CoV, yellow); Canine Coronavirus strain UCD1 (Canine CoV, hot pink); SARS-CoV-2 (SARS, brown).

### Evaluating the sensitivity of SARS-CoV-2 RBD for serology

To evaluate the sensitivity of the RBD of SARS-CoV-2 for identifying infected individuals, we obtained a total of 77 serum samples from 70 patients with laboratory confirmed (PCR positive) SARS-CoV-2 infections collected at different times after the onset of symptoms. All the samples were tested for binding of total Ig and IgM antibodies to recombinant RBD antigens from SARS-CoVs and common-cold HCoVs. The sensitivity of the assay was high (98% and 81% respectively for Ig and IgM) for specimens collected 9 days or more after onset of symptoms (Figure 4A). As expected, overall sensitivity was lower (57% and 43 % respectively for Ig and IgM) for specimens collected between 7 and 8 days after onset of symptoms (Figure 4A). With samples collected 9 days or more after onset of symptoms, we observed some Ig and IgM antibody cross reactivity with the RBD of SARS-CoV-1 (67% and 30% respectively for Ig and IgM), which was anticipated as these viruses are closely related group 2B β-coronaviruses (20, 21). When the specimens were further analyzed to estimate the timing of seroconversion, we observed a marked transition from seronegative to positive for both Ig and IgM about 9 days after the onset of symptoms (Figure 4B and C). For 14 individuals with laboratory confirmed SARS-CoV-2 infection, we had two specimens collected at different times early in the infection (Figure 4D). Two subjects (U80 and U02) were seronegative within the first 4 days and seropositive for both Ig and IgM 9 or more days after onset (Figure 4D). For three subjects (E09, E04, U04) the acute samples were collected after 9 days and the convalescent samples were collected 21 days or more after onset. In these individuals both acute and convalescent samples were positive, and we observed an increase in Ig and IgM levels in the second specimen. For the remaining 9 subjects, the acute specimen was collected on day 7 after onset and the convalescent specimen was collected >9 days after onset. Six out of the 9 subjects already had specific Ig, IgM or both in the acute specimen collected on day 7. All the subjects except one (U05) seroconverted or had elevated levels of antibody in the convalescent sample collected >9 days after onset of symptoms. These results indicate that most people seroconvert between days 7 and 9 after onset of symptoms. Subject U05 was an outlier and did not develop specific antibodies Ig or IgM antibodies. All the individuals with documented SARS-CoV-2 had Ig but not IgM antibodies that bound to the RBD of common HCoVs, which is consistent with their high prevalence in humans (Figure 4A). These results demonstrate that the RBD of SARS-CoV-2 is a highly sensitive antigen for antibody detection in patients 9 days or more after onset of symptoms.

**Figure 4.**
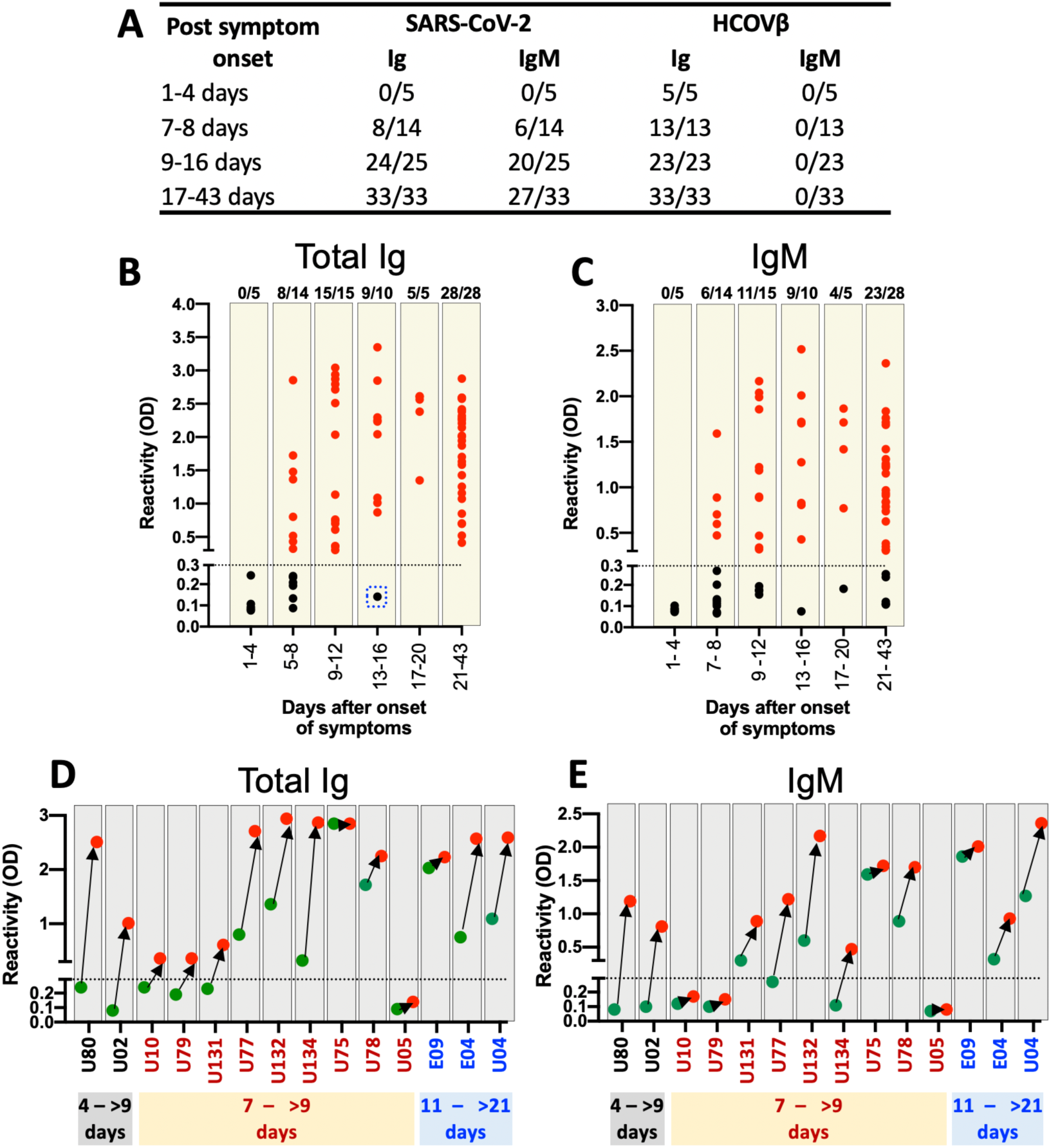
Evaluation of SARS-CoV-2 spike RBD antigen sensitivity. **A**. Overall SARS-CoV-2 spike RBD antigen sensitivity as assessed by the in-house Ig and IgM ELISA assay using clinical specimens obtained from PCR confirmed COVID-19 subjects. The changes of the levels of (B) total Ig and (C) IgM antibodies binding to RBD of the SARS-CoV-2 spike antigen. The binding of the spike RBD antigen from SARS-CoV-2 to de-identified serum samples obtained from COVID-19 positive subjects at different time points since onset of symptom are presented. The cutoff values for the ELISA assay are indicated by the broken line. Seroconversion of **D**. total Ig and **E**. IgM antibodies against RBD of the SARS-CoV-2 spike antigen among 14 representative COVID-19 patients during the acute phase since illness onset. The first sample (green) and follow-up sample (red) are connected by black arrow. The time interval between the first and follow-up sample are provided on the x-axis. The binding signals below the broken line are denoted as seronegative.

### Antibodies to the RBD of SARS-CoV-2 as a correlate of neutralizing and protective immunity

As the RBD domain of S protein is critical for viral entry, antibodies targeting this domain of SARS-CoV-2 are likely to be neutralizing and potentially protective, as is seen in cell culture and animal models for other pathogenic CoVs (19, 22). We tested a subset of SARS-CoV-2 patient immune sera for neutralizing antibodies to assess the kinetics of neutralizing antibody development and the relationship between RBD binding antibody and neutralizing antibody (Figure 5). The neutralizing antibody kinetics in patients mirrored the kinetics of RBD antibody development (Figure 5A). Among patients with confirmed SARS-CoV-2 infection, only 14% (1/7) had neutralizing antibodies within the first 7 days after onset of symptoms. By day 9 after onset, 95% (18/19), had developed neutralizing antibodies. One subject (U05) neither seroconverted for RBD antigen nor developed neutralizing antibodies to SARS-CoV-2. We observed a strong correlation between the levels of RBD binding Ig and IgM antibodies and the levels of SARS-CoV-2 neutralizing antibodies in patients (Figure 5B and 5C).

**Figure 5.**
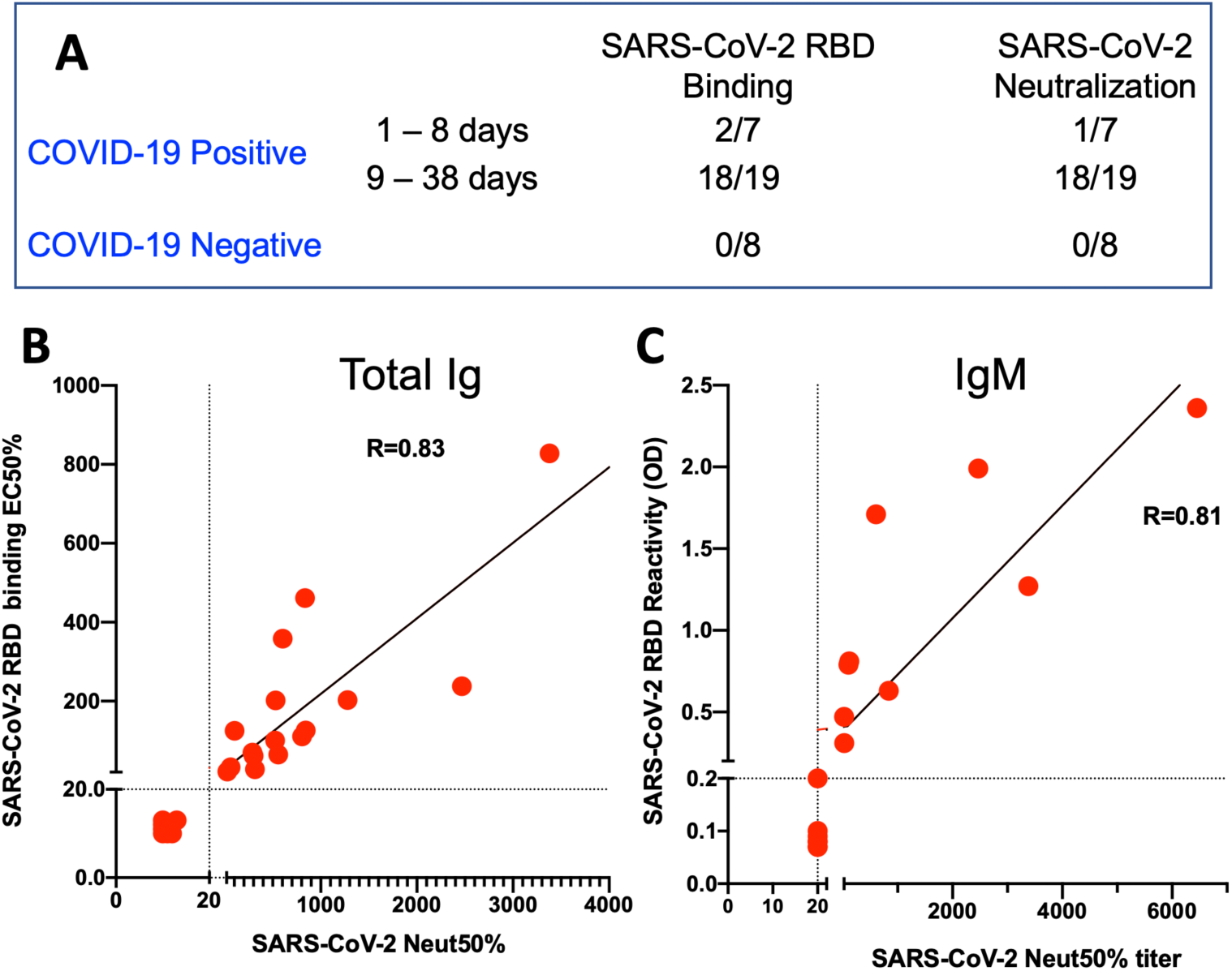
Correlation between spike RBD antigen binding and SARS-CoV-2 neutralizing antibody titers. **A**. Comparison between SARS-CoV-2 spike RBD antigen-binding ELISA and SARS-CoV-2 neutralization assay results for a total of 24 specimens (COVID-19 positives and 8 COVID-19 negatives) are presented. Correlations between **B**. total Ig and **C**. IgM RBD binding and the SARS-CoV-2 neutralizing antibody titers. A total of 23 serum samples collected between 1 and 33 days after onset of symptoms from PCR confirmed COVID-19 subjects were measured for Ig and IgM binding to spike RBD antigen and SARS-CoV-2 neutralization assay. Scatter plots were generated using individual serum binding to RBD antigen (Y-axis) versus SARS-CoV-2 neutralizing antibody titers (X-axis). Pearson Product-Moment Correlation Coefficient (R) for RBD binding and Neutralization titers are shown in the plots.

## DISCUSSION

Serology is critical to understanding the transmission, pathogenesis, mortality rate and epidemiology of emerging viruses. In the few months after the discovery of SARS-CoV-2 as a human pathogen, scientists have developed a large number of antibody assays and many tests are now available in the commercial market. Although none of the assays have been fully validated yet, the FDA has granted emergency use authorization (EUA) for a few tests, while stressing the need for further validation. Investigators have already encountered problems with the specificity and sensitivity of commercial assays rushed to market (4, 23). Widespread use of inaccurate antibody assays could lead to policies that exacerbate the current SARS-CoV-2 pandemic instead of containing it.

To address the need for reliable antibody-based diagnostic assays, we focused on the RBD domain of the spike protein because this region is poorly conserved between different CoVs and is also known to be a major target of human antibodies (19). A major concern with using a protein domain instead of a full-length protein or whole virion for antibody detection may reduce assay sensitivity. However, we observed that over 95% of SARS-CoV-2 patients develop antibodies to the RBD 9 days after onset of symptoms. We also observed specificity of well over 99%, even when using recent convalescent sera from people infected with common HCoVs or animals hyperimmunized with other zoonotic CoVs. Some patients infected with SARS-CoV-2 had antibodies that cross-reacted with the RBD of SARS-CoV-1. Since SARS-CoV-1 seroprevalence is very low in humans, the SARS-CoV-2 antibody cross-reactivity with SARS-CoV-1 is unlikely to pose diagnostic challenges. Other recent studies that have been published or under peer review also support the high specificity and sensitivity of the SARS-CoV-2 RBD for antibody detection (4, 17, 18). Amanat and colleagues tested samples from seven SARS-CoV-2 patients collected at the beginning of the epidemic in the USA and reported that the full length S protein and the RBD performed well for specific antibody detection (17). Okba and colleagues compared the performance of different SARS-CoV-2 antigens for antibody detection using samples from 10 SARS-CoV-2 patients in Europe (4). For the SARS-CoV-2 spike RBD, they observed levels of specificity and sensitivity that was comparable to our results reported here. The S2 subunit, which comprises conserved regions between CoVs, was less specific than the RBD (4). Perera and colleagues evaluated the performance of the RBD for antibody detection using samples from 24 SARS-CoV-2 patients in Hong Kong (18). They also observed overall high specificity and sensitivity when patients were tested 10 days or more after onset of illness. Our study with 77 specimens from 70 documented SARS-CoV-2 patients presenting to hospitals in North Carolina and Georgia, together with these recent studies conducted in New York, Europe and Hong Kong, strongly support the use of SARS-CoV-2 RBD as an antigen for antibody detection.

We designed the assay for separate detection of RBD-specific total immunoglobulin (Ig) and IgM. As the pandemic is ongoing and most infections are likely to have occurred within the past few months, infected individuals are likely to have variable levels of antigen-specific IgG, IgM and IgA. To maximize assay sensitivity and to prevent one antibody isotypes competing for binding sites and reducing assay signal, we measured total Ig. We did not observe any decrease in assay specificity by designing the assay to monitor levels of total Ig instead of IgG binding to the RBD (data not shown). We also tested all patient samples for IgM alone because IgM antibodies are relatively short lived and indicative of a recent exposure. When conducting large scale population level surveillance for SARS-CoV-2 antibodies, it will be possible to distinguish recent from remote infections by measuring both total specific-Ig and IgM binding to the RBD.

Antibody assays that are correlated with protective immune responses in individuals who have recovered for SARS-CoV-2 infection and also reflect herd immunity at a population level are urgently needed to define each individual’s risk of disease and to identify communities at high risk for new waves of infection. In animal studies with SARS-CoV-1, virus neutralizing antibodies were strongly correlated with protective immune responses (19). We observed a striking correlation between the levels of RBD antibodies in patients and the ability of patient sera to neutralize SARS-CoV-2 virus. Other groups have also recently reported on strong correlation between Spike/ RBD antibodies and SARS-CoV-2 neutralization patients infected with SARS-CoV-2 (4, 17, 18). While further studies are needed to fully evaluate RBD antibodies as correlate of protective immunity, the results to date indicate that RBD antibodies are a promising correlate of protection. A simple antibody detection assay that also predicts individual level risk of disease will be a major advance because SARS-CoV-2 neutralization assays are time-consuming and require BSL-3 containment.

One SARS-CoV-2 patient (U05) who tested positive for viral RNA and required hospitalization did not develop RBD-specific Ig, IgM or neutralizing antibodies even at 16 days after the onset of symptoms. This was the only person among the 68 PCR positive subjects who did not seroconvert by 9 days after onset of symptoms. While we cannot rule out the possibility of false positive PCR test result, others have also reported rare instances where people infected with SARS-CoVs have atypical, dampened immune responses (24). Further studies are needed to establish the frequency and significance of atypical antibody responses in SARS-CoV-2 patients and characterize the serological repertoire and epitopes targeted by the antibodies in convalescent sera.

As SARS-CoV-2 infections in the southeastern USA have started to increase relatively recently, the latest convalescent samples used for this study were collected within 90 days onset. In most patients, the convalescent sera had high end-point titers (>1:1000) in the RBD Ig ELISA supporting the utility of this assay even as antibody levels start to wane over time. We need to prioritize studies to prospectively monitor SARS-CoV-2 patients to determine the long-term kinetics of antibody levels and the performance antibody detection assays over time.

All the SARS-CoV-2 human immune sera used for this study were collected from symptomatic patients that included many with serious illness requiring hospitalization. The research community currently does not know if individuals experiencing inapparent/ mild symptoms after SARS-CoV-2 infection have similar kinetics and levels of RBD-binding antibodies as those experiencing symptomatic infections. Studies must be done with individuals experiencing mild/inapparent SARS-CoV-2 infections to define the kinetics and levels of RBD antibodies before implementing large population-level antibody testing.

### Experimental Methods

#### Structural analysis

The structure coordinate sets of the spike proteins, spike protein complexes with their cognate receptor ACE2 and monoclonal antibodies were obtained from the Protein Data Bank (PDB). The structures were aligned to the reference spike protein using the PyMOL Molecular Graphics System (Version 1.2r3pre, Schrödinger, LLC). Molecular figures were drawn in the PyMol. The PDB coordinates used for the structural alignments and analysis were as follows: SARS-CoV-2 spike (6VSB), SARS-CoV-1 spike (6CRV), SARS-CoV-1 spike/S230 (6NB6), SARS-Co-V1 spike RBD/80R (2GHW), SARS-CoV-1 spike RBD/ m396 (2DD8), SARS-CoV-1 spike RBD/F26G19 (3bgf), SARS-CoV-2 spike RBD/CR3022 (6W41).

#### Protein expression and purification

We used the following structure coordinates of the Coronavirus spike proteins from the PDB to define the boundaries for the design of RBD expression constructs: SARS-CoV-2 (6VSB), SARS-CoV-1 (6CRV), HKU-1 (5I08), OC43 (6NZK), 229E (6U7H) NL63 (6SZS). Accordingly, a codon-optimized gene encoding for S1-RBD [SARS-CoV-1 (318 – 514 aa, P59594), SARS-CoV-2 (331 – 528 aa, QIS60558.1), OC43 (329 – 613 aa, P36334.1), HKU-1 (310 – 611 aa, Q0ZME7.1), 229E (295 – 433 aa, P15423.1) and NL63 (480 – 617 aa, Q6Q1S2.1)] containing human serum albumin secretion signal sequence, three purification tags (6xHistidine tag, Halo tag, and TwinStrep tag) and two TEV protease cleavage sites was cloned into the mammalian expression vector pαH. S1 RBDs were expressed in Expi293 cells (Thermofisher) and purified from the culture supernatant by nickel-nitrilotriacetic acid agarose (Qiagen).

#### Generation of SARS-CoV-2 Spike VRP and immunized mouse sera

To generate virus replicon particles (VRPs), the SARS-CoV-2 S gene was inserted into pVR21 3526 as previously described (25). In summary, the SARS-CoV-2 S gene was ligated into pVR21 following digestion by restriction endonuclease sites, Pac1 and Apa1. T7 RNA transcripts were generated using the SARS-CoV-2-S-pVR21 construct in conjunction with plasmids containing the VEEV envelope glycoproteins and capsid protein. The RNA transcripts were then electroporated into BHK cells and monitored for CPE. VRP were harvested 48 hours after electroporation and purified via high-speed ultra-centrifugation. To generate serum samples against SARS-CoV-2, 10-week-old BALB/c mice (Jackson Labs) were inoculated via footpad injection with the VRP and boosted with the same dose one time three weeks later. Serum samples were then collected from individual animals at 2 weeks post-boost and pooled for use in assays.

#### Human specimens

All human specimens used in these studies were obtained after informed consent under good clinical research practices (GCP) and compliant with oversight by the relevant institutional review boards (IRBs).

UNC Hospital Specimens: Sera for this study were remnants from samples submitted to the UNC Hospital McLendon Clinical Laboratories or Blood Bank. SARS-CoV-2 positive patient samples were obtained from patients with positive RT-PCR test result for SARS-CoV-2. SARS-CoV-2 negative samples were obtained from patients with other diagnoses or from samples collected prior to December 2019 and cryopreserved at −80·C.

Emory University School of Medicine Specimens: Specimens were obtained from patients with symptomatic illness and clinical testing confirming SARS-CoV-2 by PCR. De-identified specimens were shared with researchers at UNC consistent with local IRB protocols (Emory IRB# 00110683 and 00022371).

Healthy Unexposed Donors: Samples from healthy US adult donors were obtained by the La Jolla Institute for Immunology (LJI) Clinical Core or provided by a commercial vendor (Carter Blood Care) for prior, unrelated studies between early 2015 and early 2018, at least one year before the emergence of SARS-CoV-2. The LJI Institutional Review Board approved the collection of these samples (LJI; VD-112). Samples from the Caribbean, Central America and South Asia were were obtained from archived samples at UNC collected before December 2019 for other studies.

Human and Animal Specimens from BEI resources: The following reagent was obtained through BEI Resources, NIAID, NIH as part of the Human Microbiome Project: Pooled sera obtained from rabbits dosed with a recombinant SARS-CoV spike protein (NRC-772), monoclonal anti-SARS-CoV S protein (Similar to 240C) (NR-616), anti-porcine respiratory coronavirus (PRCoV; ISU-1) serum obtained from Pig (NR-460), anti-porcine Transmissible Gastroenteritis Virus obtained from Pig (NR-458), anti-porcine respiratory coronavirus (PRCoV; ISU-1) serum obtained from Guinea Pig (NR-459), Anti-SARS Coronavirus obtained from Guinea Pig (NR-10361), Anti-Bovine Coronavirus (mebus) obtained from Guinea Pig (NR-455), Anti-Feline Infectious Peritonitis Virus, 79-1146 obtained from Guinea Pig (NR-2518), Anti-Avian Infectious Bronchitis Virus, Massachusetts obtained from Guinea Pig (NR-2515), Anti-Turkey Coronavirus, Indiana obtained from Guinea Pig (NR-9465), Anti-Canine Coronavirus, UCD1 obtained from Guinea Pig (NR-2727), Anti-Human Parainfluenza Virus 2 obtained from Guinea Pig (NR-3231), Anti-Simian Virus 5 obtained from Guinea Pig (NR-3232), Anti-Human Parainfluenza Virus 3 obtained from Guinea Pig (NR-3235), Anti-Bovine Parainfluenza Virus 3 obtained from Guinea Pig (NR-3236), Anti-Human Parainfluenza Virus 4A obtained from Guinea Pig (NR-3239), Anti-Human Parainfluenza Virus 4B obtained from Guinea Pig (NR-3240), Human Convalescent Serum 001 to 2009 H1N1 Influenza A Virus (NR-18964), Human Convalescent Serum 002 to 2009 H1N1 Influenza A Virus (NR-18965), and Human Reference Antiserum to Respiratory Syncytial Virus Human respiratory syncytial virus (NR-4020).

#### *In-house* RBD Ig and IgM ELISA

All serum specimens tested by ELISA assay were heat-inactivated at 56°C for 30 minutes to reduce risk from any possible residual virus in serum. Briefly, 50 μl of spike RBD antigen at 4 μg/ml in Tris Buffered Saline (TBS) pH 7.4 was coated in the 96-well high-binding microtiter plate (Greiner bio one cat # 655061) for 1 hr at 37°C. Then the plate was washed three times with 200 μl of wash buffer (TBS containing 0.2% Tween 20) and blocked with 100 μl of blocking solution (3% milk in TBS containing 0.05% Tween 20) for 1 hr at 37°C. The blocking solution was removed, and 50 μl of serum sample at 1:20 or indicated dilutions in blocking buffer was added for 1 hr at 37°C. The plate was washed in the wash buffer, 50 μl of alkaline phosphatase-conjugated secondary goat anti-human secondary Ab at 1:2500 dilution was added for 1 hr at 37°C 1. For measuring total Ig, a mixture of anti-IgG (Sigma Cat # A9544), anti-IgA (Ab cam Cat # AB97212), and anti-IgM (Sigma Cat # A3437] were added together. For measuring IgM, only goat anti-human IgM was used. The plate was washed, and 50 μl P-Nitrophenyl phosphate substrate (SIGMA FAST, Cat No N2770) was added to the plate and measured absorbance at 405nm using a plate reader (Biotek Epoh, Model # 3296573). For testing animal sera, the secondary antibody was matched to the species as follows: goat anti-mouse IgG (Sigma, A3688), goat anti-rabbit IgG (Abcam, ab6722), goat anti-pig IgG (Abcam, ab6916), and goat anti-guinea pig IgG (Abcam, ab7140).

#### SARS-CoV-2-Washington neutralization assays

Full-length viruses expressing luciferase were designed and recovered via reverse genetics and described previously (26, 27). Viruses were tittered in Vero E6 USAMRID cells to obtain a relative light units (RLU) signal of at least 20X the cell only control background. Vero E6 USAMRID cells were plated at 20,000 cells per well the day prior in clear bottom black walled 96-well plates (Corning 3904). Neutralizing antibody serum samples were tested at a starting dilution of 1:20, and were serially diluted 4-fold up to eight dilution spots. Antibody-virus complexes were incubated at 37°C with 5% CO_2_ for 1 hour. Following incubation, growth media was removed and virus-antibody dilution complexes were added to the cells in duplicate. Virus-only controls and cell-only controls were included in each neutralization assay plate. Following infection, plates were incubated at 37°C with 5% CO_2_ for 48 hours. After the 48 hour incubation, cells were lysed and luciferase activity was measured via Nano-Glo Luciferase Assay System (Promega) according to the manufacturer specifications. SARS-CoV-2 neutralization titers were defined as the sample dilution at which a 50% reduction in RLU was observed relative to the average of the virus control wells.

## Data Availability

Antigens are available under an MTA. Please contact Lakshmanane Premkumar or Aravinda M. de Silva

## Acknowledgements

We gratefully acknowledge BEI (https://www.beiresources.org) resources for the prompt processing and shipping of the reagents. We are grateful for the expert procedural care provided by the UNC Hospital, Blood Donor Center and to patients and blood donors providing samples for the study. This work was funded by the University of North Carolina School of Medicine (PL and AD), National Institutes of Health contract Nr. 75N9301900065 (A.S. and D.W.), NIH NIAID T32 AI007151 (DM) and a Burroughs Welcome Fund Postdoctoral Enrichment Program Award (DM).

